# Unpicking the Gordian knot: Mendelian randomization to elucidate the risk factors for infectious diseases, using EBV as a model pathogen

**DOI:** 10.1101/2022.02.04.22270455

**Authors:** Marisa D. Muckian, James. F Wilson, Graham S. Taylor, Helen R. Stagg, Nicola Pirastu

## Abstract

**Background:** Why particular individuals are more at risk of a given infectious disease than others has been a topic of interest for scientists, clinicians, and polymaths for millennia. Complex webs of factors-sociodemographic, clinical, genetic, environmental-intersect, rendering causality difficult to decipher. We aimed to demonstrate the ability of Mendelian Randomization (MR) to overcome the issues posed by confounding and reverse causality to determine the causal risk factors for the acquisition of infectious diseases, using Epstein Barr Virus (EBV) as a model pathogen.

**Methods:** We mapped the complex evidence from the literature prior to this study factors associated with EBV serostatus (as a proxy for infection) into a causal diagram to determine putative risk factors for our study. Using data from the UK Biobank of 8,422 individuals genomically deemed to be of white British ancestry between the ages of 40 and 69 at recruitment between the years 2006 and 2010, we performed a genome wide association study (GWAS) of EBV serostatus, followed by a Two Sample MR to determine which putative risk factors were causal.

**Results:** Our GWAS identified two novel loci associated with EBV serostatus. In MR analyses, we confirmed educational attainment, number of sexual partners, and smoking as causal risk factors for EBV serostatus.

**Conclusions:** Our study demonstrates the power of MR to decipher complex webs of putative risk factors and determine which are causal for the acquisition of an infectious disease. The factors identified for EBV will be important for vaccine deployment.

**Key messages:**

- The risk of infectious disease acquisition is dependent on many interacting sociodemographic, lifestyle, clinical, genetic, environmental, and national and international health governance factors.
- Traditional epidemiological studies of these risk factors are often hindered by issues of confounding and therefore whether a given putative risk factor is causally associated with infection acquisition is difficult to decipher.
- Using Epstein Barr Virus (EBV) as a model pathogen, we demonstrate the power of Mendelian randomization to understand if putative risk factors are causal, while controlling for confounding.
- Better understanding of infectious disease risk factors using Mendelian randomization can inform vaccine strategies and deployment e.g. by identifying priority populations for vaccination.

## Introduction

The risk of acquiring an infectious disease is dependent on an interacting mix of factors spanning from sociodemographic to lifestyle, clinical, genetic, environmental, and national and international health governance. For example, tuberculosis (TB) disease has a range of interacting factors and comorbidities such as infection with human immunodeficiency virus (HIV), overcrowding, malnutrition, smoking, diabetes, and alcohol use.^1^ Some of these risk factors are common across many infectious diseases, but some are more specific to particular infections, such as lifestyle factors which include exposure to infection in a professional environment (e.g. schistosomiasis), recreational drug use (e.g. blood borne viruses), and sexual behaviour (e.g. herpes simplex virus 2, HSV-2).^2^ Clinical factors such as transplantation and immunodeficiencies increase the risk for opportunistic infections such as cytomegalovirus (CMV).^2,33^ Traditional epidemiological studies are limited in their ability to pull apart such complex webs of evidence to determine actual causality and the relative contribution of different causal factors. They are often impacted by unmeasured confounding and the possibility of reverse causality.

Mendelian randomisation (MR) is a technique that takes advantage of genetic data to understand if a putative risk factor of interest is causally associated with a given health outcome. Unlike traditional epidemiological studies, MR eliminates the issues of unmeasured confounding and reverse causality using instrumental variables (IVs), which are genetic variants known to be associated with the risk factor. This means it is possible to accurately determine if a putative risk factor is causally associated with an outcome, provided the assumptions of MR are met.

Epstein Barr Virus (EBV) is a human herpes virus infecting ∼90% of the global population and is a pathogen for which the evidence on the risk factors for infection is complex. It is associated with 164,000 cancer deaths per year^4^ as well as multiple sclerosis.^5^ The burden of disease associated with EBV is such that interest in the development of infection-or disease-preventing vaccines is extensive. A Phase II trial of an early vaccine candidate only reduced symptom severity upon infection,^6^ the development of more immunogenic candidates^7^ and the success of the Pfizer/BioNTech and Moderna mRNA vaccines against SARS-CoV-2 has given impetus to EBV prophylactic vaccination. Indeed, Moderna currently have a mRNA-based EBV vaccine candidate in clinical development.^8^ Knowledge on the risk factors for EBV infection is critical to determine the best model for infection-preventing vaccine deployment and to understand why some individuals remain EBV negative for life, which is informative to the consequences of ‘induced’ non-infection with EBV through vaccination.

Extensive work has been undertaken internationally to determine the risk factors for EBV infection. As highlighted in a recent systematic review,^9^ research to date has focussed on sociodemographic, dietary, and lifestyle factors. A small number of studies have also examined genetic susceptibility to EBV infection.^10–16^ Recent studies have identified genetic variants associated with anti-EBV antibody levels ^12,13,17^Although some risk factors are consistently displayed from setting to setting (age being the clearest example), the published population health literature is often contradictory. This is due in part to poor control for confounding in such studies, partly due to the cost and complexity of measuring all putative relevant factors simultaneously. MR provides an opportunity to untie this Gordian knot.

This study sought to demonstrate the value of MR in determining the risk factors for the acquisition of infectious diseases, using EBV as a model infection. We performed a genome-wide associated study (GWAS) to determine the genetic risk factors for EBV infection, followed by an MR to interrogate the published putative non-genetic factors, all within the UK Biobank (UKB), a UK based cohort study of people aged between 40-69 years.^18^ Our study demonstrates the power of MR in overcoming the pitfalls of traditional epidemiological approaches, not only for EBV, but also for other infectious diseases.

## Methods

To perform an MR on the association between the acquisition of EBV infection and different putative risk factors, we undertook the following steps, each of which are laid out in separate sections of the methods. 1) Identify a population of interest for the analysis within which 2) EBV serostatus (as a proxy for infection) had been tested for and 3) which had been genotyped. 4) Identify the putative risk factors of interest for EBV infection from the published literature. 5) Descriptively analyse the population of interest in light of the putative risk factors of interest. 6) Find corresponding existing GWASs to extract instrumental variables (genetic variants known to be associated our putative risk factors of interest. 7) Undertake a GWAS of EBV serostatus and where pre-existing GWASs could not be found for a putative risk factor of interest. 8) Perform MR.

### Study population

UKB is a prospective cohort study of over 500,000 participants recruited in the UK between 2006-2010. Participants of the UKB were aged between 40 and 69 years old at the time of recruitment.^18^

### Epstein Barr Virus serostatus

A subset of 9,695 participants in the UKB were subject to serological testing on samples taken at the point of their enrolment into the cohort, including for anti-EBV antibodies. A multiplex serology-based approach, as described by Brenner *et al*., ^19^ was used for testing. Antibody levels against different EBV antigen targets were expressed as median fluorescence intensity (MFI). (Data were recorded by the UKB as both antibody levels against each antigen and in a binary format (seropositive/seronegative) for overall EBV serostatus if two or more MFI thresholds were met. As EBV is a herpesvirus that establishes a lifelong infection in humans, we used serostatus as a proxy for EBV infection throughout our analyses.

### Genotyping

UKB participants had DNA extracted from samples taken during their initial visit to one of 22 assessment centres. Genotyping was carried out using the Applied Biosystems UK Biobank Axiom Array and the UK BiLEVE array. Autosomal single nucleotide polymorphisms (SNPs) were imputed using a merged reference panel of the Phase 3 1000 Genome Project and UK10K using IMPUTE3. Procedures are described in full by Bycroft *et. al*.^18^

### Exposures to be tested for causal effect on EBV serostatus

Non-genetic variables to be tested for a putatively causal effect on EBV serostatus through MR were selected based on a review by Winter *et. al*,^9^ Six factors (childhood household size, total number of sexual partners, BMI, tonsillectomy, educational attainment, smoking status) were selected on the basis of the balance of evidence within the review being in favour of a putative causal effect (Supplementary table 1) and mapped into a causal diagram (Figure 1) in broad groups: household size, lifestyle factors (smoking status), socioeconomic factors (educational attainment), genetic factors, clinical factors (BMI, tonsillectomy). The review found coinfection with other several other viruses to be associated with EBV status including: human immunodeficiency virus (HIV), Kaposi’s sarcoma related herpes virus (KSHV), human T-lymphotropic virus (HTLV), CMV, and herpes-simplex 1 (HSV-1). We did not include these in our MR studies due to potential of overlapping genetic variants that may influence both the risk factor infection and EBV.

**Figure 1:**
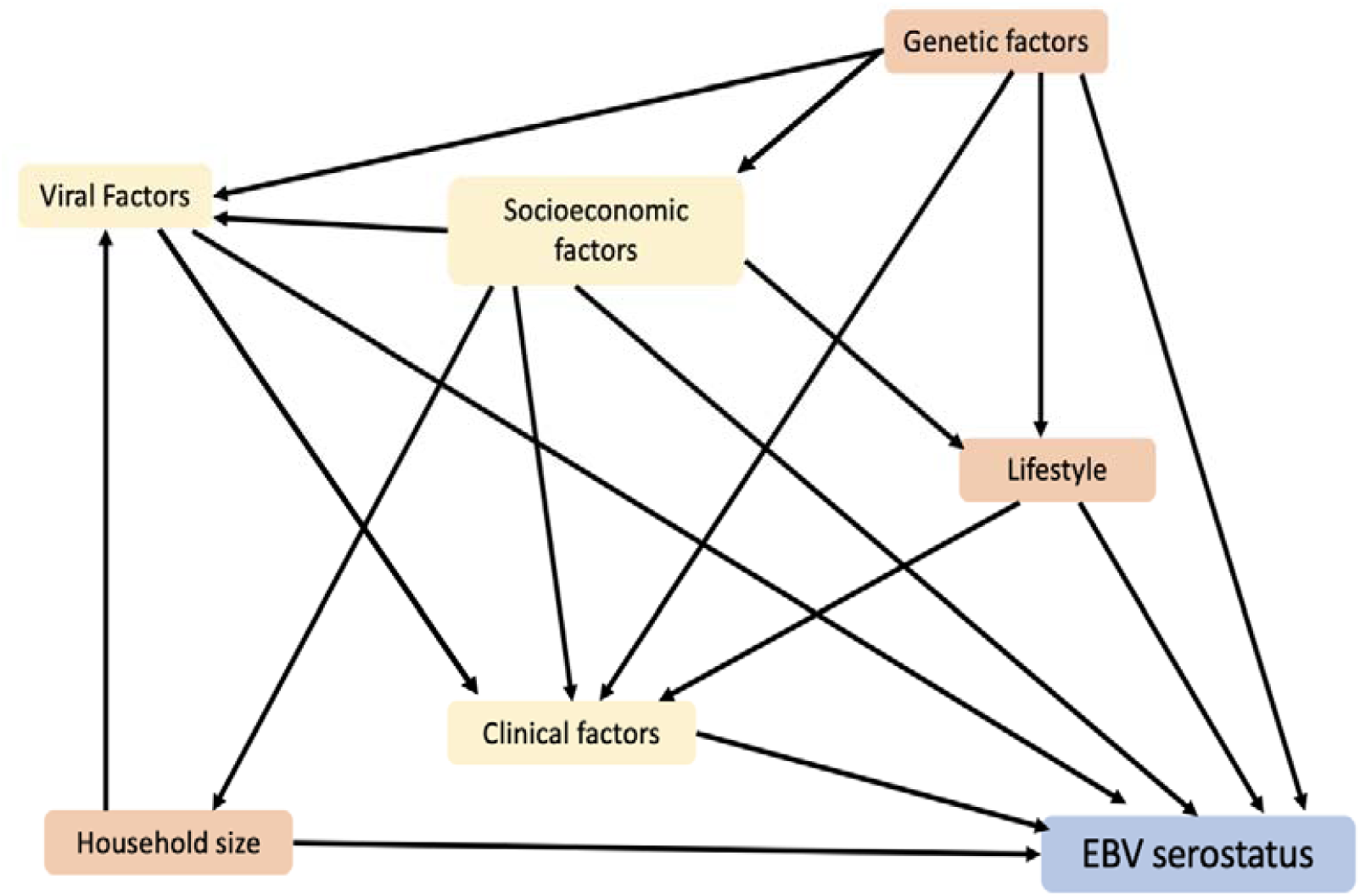
Causal diagram of risk factors for EBV infection. Diagram contains risk factors mapped to broad categories such as household size, lifestyle factors (smoking status), socioeconomic factors (educational attainment), and clinical factors (body mass index, tonsillectomy)

### Descriptive analysis

A description of the demographics of the overall UKB cohort and the subcohort of individuals with EBV serostatus and who were genomically deemed to be of white British ancestry (see genome-wide association study section of the methods) was carried out using R (v3·6·1).^20^ Qualitative traits are reported as a % total (N). Normality of the quantitative variables was assessed using a Kolmogorov-Smirnov test and non-normally distributed traits were subsequently expressed using the median and interquartile range.

### Exposure instrumental variable selection

Next, IVs for each exposure (genetic variants associated with the exposures i.e., putative risk factors, in this case, SNPs) were selected. From the causal diagram, six putative risk factors were selected. For all risk factors apart from household size, IVs were obtained from previously published GWAS results (Supplementary table 2). For each exposure variable, the largest and most recent GWAS performed in European samples was used. From each exposure GWAS we selected genome wide significant (*p*<5×10^−8^) and independent SNPs (r^2^<0·0001) using the TwoSampleMR package. For total number of siblings GWAS results were not available, thus we performed our own GWAS using the individuals from UKB who did not participate in the serological study (N=319,209). For total number of siblings, we combined the total number of sisters and total number of brothers variables as reported in the questionnaire of UKB. GWAS methods are described below.

### Genome-wide association study for EBV serostatus

As a preparatory step for the MR, we carried out two GWAS, 1) of EBV serostatus on the subcohort, our MR outcome variable and 2) of household size as measured by total number of siblings, as no IV could be identified from the literature for this putative risk factor. These were carried out on UKB participants with genomic data, including only unrelated individuals and those who were genomically deemed to be of white British ancestry determined using a principal component analysis (PCA) performed by UKB.^18^ Analysis was carried out using an in-house GWAS pipeline employing a two-step GRAMMAR-Gamma framework. The two phenotypes were regressed against the fixed effect covariates (sex, age, genotyping batch, array type, and the first 40 principal components (PCs) as calculated by UKB to account for population substructure).^21^ Fixed effect residuals were then further corrected for the effect of relatedness by using FastGWA^22^ which corrects the trait based on the sparse genetic relatedness matrix (GRM), creating the GRAMMAR-Gamma residuals to be used for the association analysis. In step 2, these GRAMMAR-Gamma residuals were regressed against genome-wide SNP dosages using RegScan.^23^ Genome-wide association was performed using a linear regression model. Genome-wide significant loci were defined using a p-value threshold of 5×10^−8^. The resulting SNPs from the total number of siblings GWAS were chosen as IVs for MR analysis and included independent significant SNPs (r^2^=0·001, *p*= 5×10^−8^).

### Mendelian randomization

MR analysis allows us to determine the causal role that a given exposure (X) has on a given outcome (Y) without the impact of confounding. Genetic instruments-such as SNPs-that directly affect the exposure of interest, can be used as IVs to determine the exposure’s effect on a specific outcome. If individuals with genetic variants associated with the risk factor, have a higher incidence of the outcome, in this case EBV seropositivity, we can conclude that the risk factor is causal for EBV. Genetic variants are valid instruments so long as they are not also associated with the outcome and are not influenced by any confounders (U) (Figure 2).

**Figure 2:**
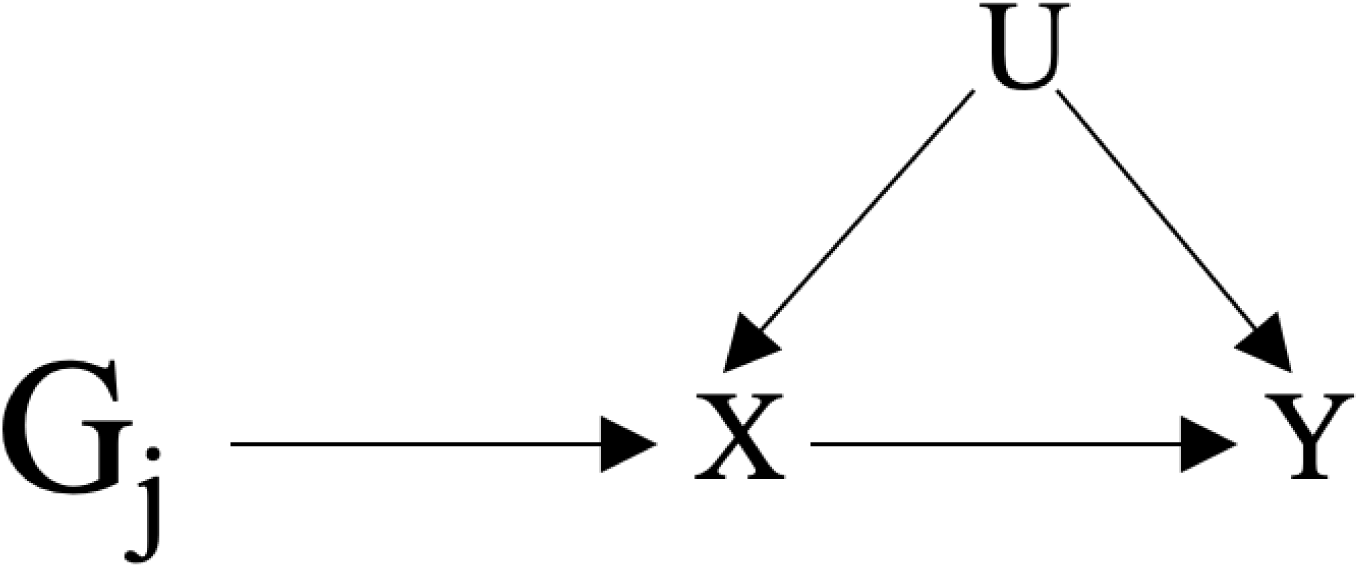
Mendelian Randomization. Mendelian randomization uses genetic instruments (G_j_) associated with the exposure (X) of interest as instrumental variables, to determine the causal relationship of X on the outcome (Y) without the influence of confounding. The instruments must not be association with any confounders (U).

For this study we used Two-Sample MR, in which the effects of the SNP on the exposure and the outcome are estimated in two distinct set of samples. The two effect sizes are harmonized, to ensure both the outcome and exposure effects align to the same allele. The effect of the exposure on the outcome is then estimated. This method was used to test if seven (including two measures of smoking status, age at smoking initiation and ever smoking) previously identified putative risk factors had a role in influencing risk of EBV infection. As an outcome, we used our GWAS of EBV serostatus. The exposure and outcome data were harmonised before MR was performed. We firstly detected outliers using the RadialMR package and IVW radial function. We then removed these outliers from the harmonised data. To test the validity of our causal inferences we carried out multiple sensitivity analyses. Firstly, using TwoSampleMR, we calculated Cochran’s Q statistic to assess heterogeneity of the genetic instruments. We then checked for directional pleiotropy using the Egger regression function. To ensure no single genetic variant was impacting the causal estimate of our results, we performed a leave one out analysis using TwoSampleMR.

Multivariable MR (MVMR) was planned to assess for correlation of significant factors at the univariable stage using the Mendelian Randomization package.^24^

## Results

### Descriptive analysis

Of the 9,695 individuals within the UKB sub-cohort that underwent serological testing, 8,244 (97·3%) had available results for EBV serostatus and were genomically deemed to be of white British ancestry (Supplementary table 3, which also compares the subcohort the overall UKB cohort). Of those 7,795 (94·6%) were EBV seropositive. The age and sex distribution within the sub-cohort were like that of the main cohort.

### Genome-Wide Association Study

GWAS of EBV serostatus (positive or negative) revealed two independent genome-wide significant loci for EBV serostatus (*p*=< 5×10^−8^) (Supplementary figure 1, Supplementary table 4). The first locus was located on chromosome 13 and mapped nearest to *RASA3*, (rs71449058, *p*=2·34×10^−10^); effect allele C has a protective effect against EBV. The second locus was on chromosome 6 and the nearest gene *PREP* (rs1210063, *p*=4·01×10^−9^), the effect allele G was found to increase susceptibility to EBV.

### Mendelian randomization

Using our GWAS results, we next sought to determine if our putative risk factors for EBV serostatus were, in fact, causal. After removal of outlying IVs (Supplementary table 5), univariable MR showed that educational attainment (*p*=7·20×10^−6^), sexual partners (*p*=0·02), and smoking (*p*=0·049) were found to be associated with EBV serostatus (Table 1). For each additional year of genetically predicted education (baseline 0 years) the odds of being EBV seropositive decreased (OR=0·43, 95% CI=0·30-0·62). Compared to previous studies this we observed the opposite direction of effect (Figure 3) although effect size was difficult to compare due to differences in exposure measurements. Increase in total number of sexual partners from <2 partners to between 2-5, increased the odds of being EBV seropositive increased to 2·69 (95% CI = 1·15-6·32), consistent with previous literature. Finally, being a smoker (previous or current) increased the odds of EBV 2·36 times (95% CI=1·00-5·55). No other putative risk factors were found to be associated with EBV serostatus.

**Table 1.**
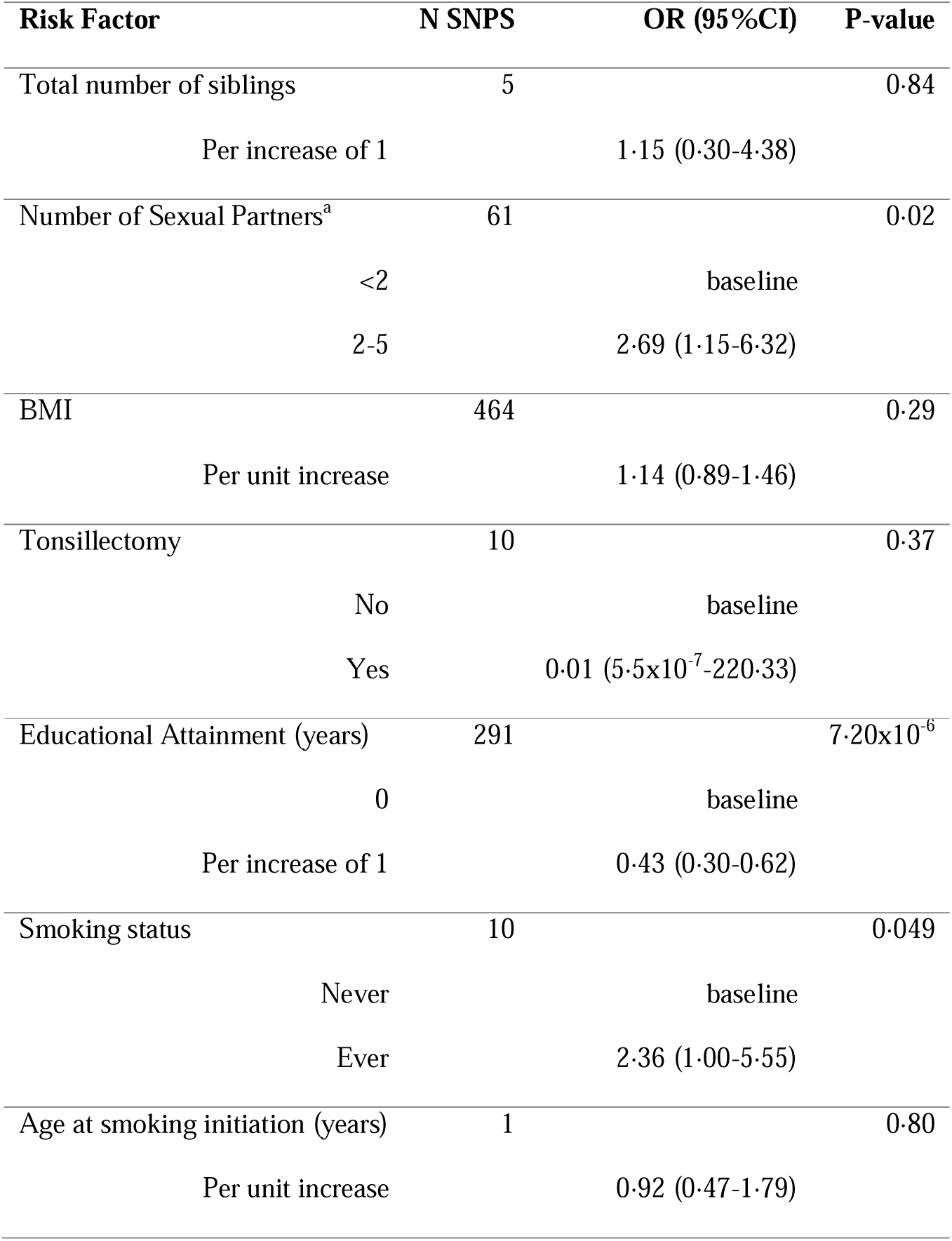
Univariable Mendelian randomization of putative risk factors for EBV infection. Putative risk factors include, total number of siblings, total number sexual partners (<2, 2-5, ≥5) ^a^(OR only calculated for <2 and 2-5), BMI, Tonsillectomy (yes/no), Educational attainment (years), smoking status (never/ever), age at smoking initiation (years). Risk factors with *p*=<0·05 were considered significant. Abbreviations: BMI-body mass index, CI – confidence intervals, N-number, OR-odds ratio, SNPs-single nucleotide polymorphisms

| **Risk Factor** | **N SNPS** | **OR (95%CI)** | **P-value** |
| --- | --- | --- | --- |
| Total number of siblings | 5 |  | 0.84 |
| Per increase of 1 |  | 1.15 (0.30-4.38) |  |
| Number of Sexual Partners<sup>a</sup> | 61 |  | 0.02 |
| <2 |  | baseline |  |
| 2-5 |  | 2.69 (1.15-6.32) |  |
| BMI | 464 |  | 0.29 |
| Per unit increase |  | 1.14 (0.89-1.46) |  |
| Tonsillectomy | 10 |  | 0.37 |
| No |  | baseline |  |
| Yes | | 0.01 ($5.5 \times 10^{-7}$-220.33) | |
| Educational Attainment (years) | 291 | | $7.20 \times 10^{-6}$ |
| 0 |  | baseline |  |
| Per increase of 1 |  | 0.43 (0.30-0.62) |  |
| Smoking status | 10 |  | 0.049 |
| Never |  | baseline |  |
| Ever |  | 2.36 (1.00-5.55) |  |
| Age at smoking initiation (years) | 1 |  | 0.80 |
| Per unit increase |  | 0.92 (0.47-1.79) |  |

**Figure 3:**
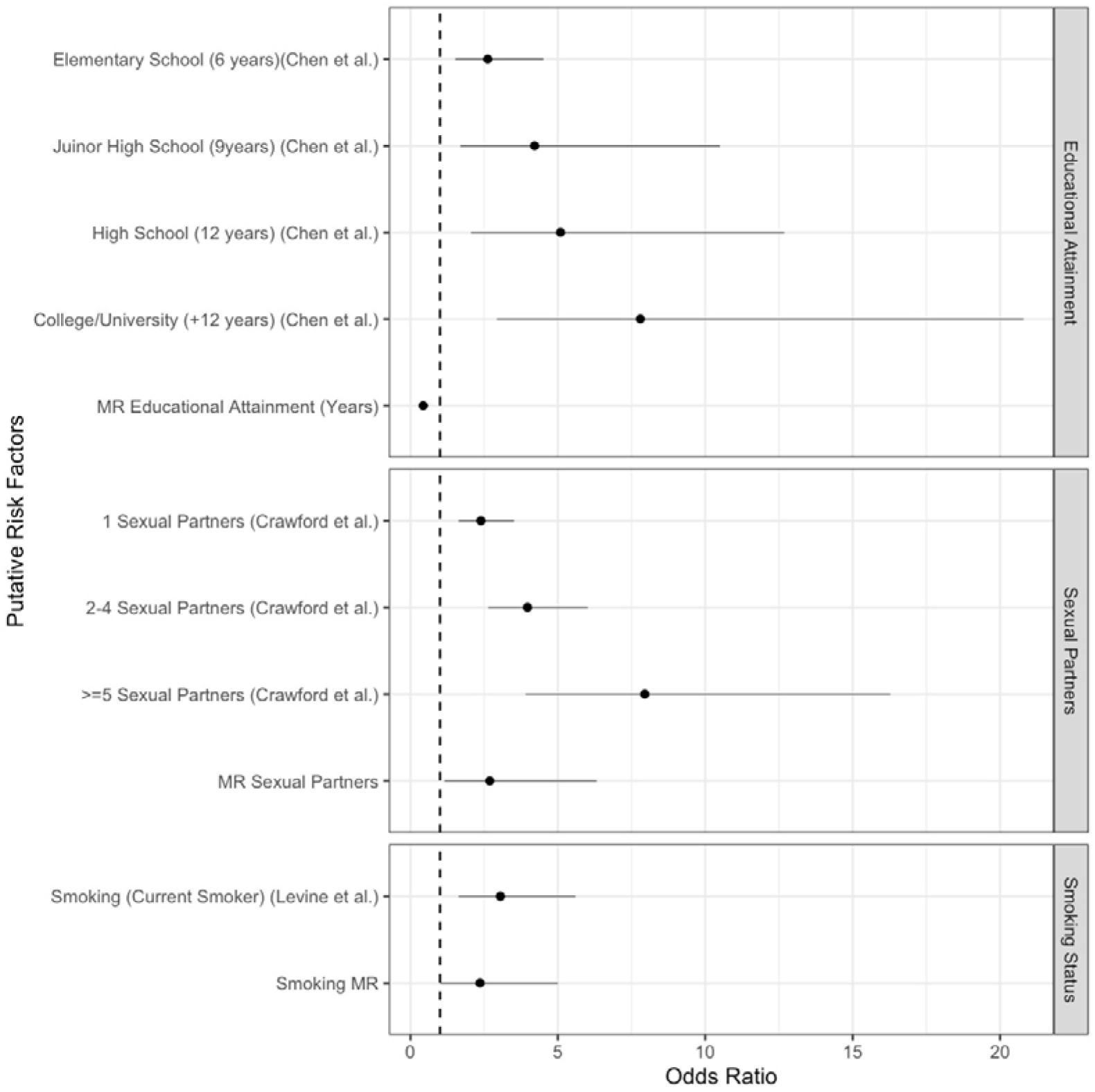
Mendelian Randomization results compared to previous observational studies for educational attainment, number of sexual partners and smoking status. Our educational attainment MR compared to an observational study in Taiwan (Baseline = uneducated) by Chen *et al*. ^28^ The sexual partners MR compared to observational effects converted from risk factors from a previous study by Crawford *et al*.^*29*^ (Baseline = 0). Smoking status MR compared to a study by Levine *et al*.^*30*^ (Baseline = never smoked). Abbreviations MR – Mendelian randomization

Sensitivity analyses demonstrated no significant heterogeneity between the estimate from each of the exposure IVs and EBV status while we detected no sign of directional pleiotropy when tested using Egger regression. Finally leave-one-out analysis showed the observed effect was constant and not driven by any single SNP (Supplementary figure 2a-c).

### Multivariable Mendelian Randomization

To determine if education, total number of sexual partners and smoking were independent risk factors for EBV we performed MVMR (Table 2). Results indicated that educational attainment was an independent risk factor for EBV (OR=0·46, 95% CI=0·32-0·67, *p*=3×10^−6^). Smoking also was an independent risk factor (OR=4·13, 95% CI=1·51-11·30, *p*=0·006). The total number of sexual partners had a similar OR to the univariable analysis (OR = 2·12, 95% CI= (0·66-6·82), but the result was not statistically significant (*p*=0·206). This could be due to fewer IVs in the MVMR being associated with total number of sexual partners, reducing power.

**Table 2.**
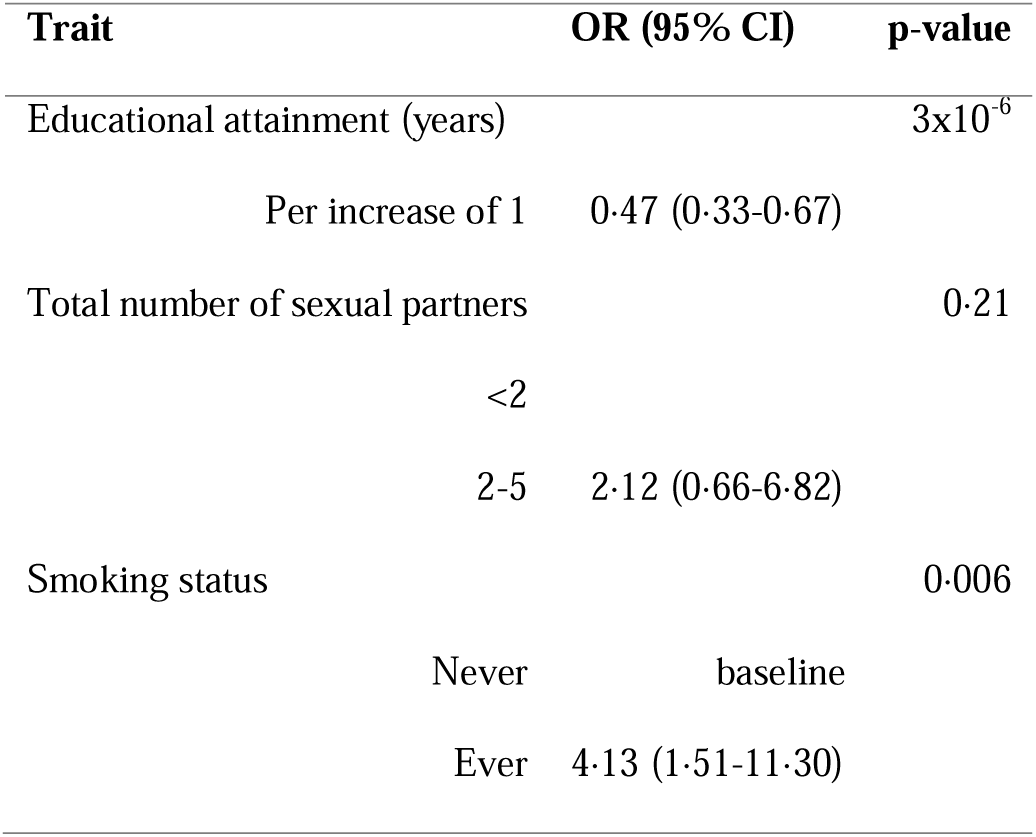
Multivariable mendelian randomization of risk factors for EBV infection. Model adjusted for three risk factors, total number sexual partners (<2, 2-5, ≥5), Educational attainment (years), smoking status (yes/no), Risk factors with p=<0·05 were considered significant. Abbreviations: CI-confidence intervals, OR-odds ratio.

## Discussion

We present the first MR to examine the causality of potential risk factors for the acquisition of an infection, using EBV as a model condition of interest. Our MR analysis of previously identified risk factors and EBV serostatus demonstrates how MR can be used to unpick the at times conflicting evidence on the complex spectrum of factors that pose a risk for the acquisition of an infectious disease. This is not only the case for EBV, but it also provides a proof of principle for other infectious diseases. In our study we identified two loci (rs1210063 and rs71449058) associated with EBV infection through an initial GWAS and provided evidence that the non-genetic factors educational attainment and number of sexual partners and smoking are likely causally associated with infection.

Examining the loci documented within our GWAS first, previous publications have documented that one of the nearest genes to these loci have previously been discussed in the EBV literature (*RASA3)*. This gene locates near viral protein binding sites that may enhance regulation of the EBV lytic cycle.^25^

Comparing our findings to previous studies of the genetics of EBV infection, it is interesting to note that such studies have focussed primarily on antibody levels. Anti-EBNA-1 levels have been established to associate strongly with the HLA class II region^12–15,17^ and more recently this region was found to be associated with anti-VCA IgG.^17^ In a recent publication, Butler-Laporte *et al*.found similar results to our GWAS, despite slight differences in sample selection, the top SNP for EBV seropositivity documented in that publication-rs71437272-showed a similarly strong result in our analysis.^16^

While our GWAS results provided insight into the genetic susceptibility component of our causal framework, they also gave us the tools required to untangle the conflicted evidence reported in the literature for EBV risk factors. An increased number of sexual partners and either being or having been a smoker increased risk of EBV. We found that having a higher educational attainment was protective for EBV in univariable MR, in contrast to the results of Chen *et al*.,^26^ possibly due to differential access to education at the relevant time points in the UK and Taiwan. Given the association in the UK between years spent in education and socioeconomic status, as well as smoking and socioeconomic status, these two findings correlate within our MR. MVMR found smoking and educational attainment to be independent risk factors for EBV status. The direction of the effect for smoking was consistent with the previous literature.^27–29^ In contrast, BMI, age at smoking initiation, total number of siblings, and having your tonsils removed were not associated with EBV in this MR analysis.

With the recent surge in interest in EBV infection-preventing vaccines, our results present an insight into the future deployment of such products based on known risk factors for EBV infection. For example, the cost of a future vaccine may limit publicly funded deployment to at-risk groups from EBV associated diseases. There is a known association between EBV acquisition at later life stages, infectious mononucleosis and then cancer,^30^ as well as a likely strong association between the time point of acquisition and population level socioeconomic status.^9^ Thus our documentation of two individual level socioeconomically associated factors (smoking^31^ and years in education) as likely causally associated with infection demonstrates an opportunity for targeted deployment of the vaccine to particular population groups. Whilst it is not possible to deploy a vaccine on the basis of a factor such as smoking status, doing so on the basis of enrolment in different levels of education is commonly used for other infectious diseases e.g. meningitis A, C, W, Y.

The core strength of our study is its demonstration of the power of MR in unpicking the complex knots of causality for the risk factors for an infectious disease. Our study population was restricted to individuals genomically deemed to be of white British ancestry, limiting generalisability. EBV seroprevalence and the age by which seroprevalence reaches equilibrium varies between populations^9^ and both genetic and non-genetic factors are likely to vary too. Additional studies across populations of different ancestries are required. Data were only available on EBV serostatus at baseline within the UKB, limiting our ability to examine risk factors in temporal proximity to EBV acquisition. UKB, like many population cohorts, is known to not be truly representative of the general population and is particularly enriched for individuals of higher educational status. Finally, our study had limited power due to 95% of individuals being EBV seropositive.

Despite these limitations, we show that MR is a powerful tool when investigating epidemiological risk factors for the acquisition of infectious diseases. Our results define a core set of factors that should be adjusted for in analyses of the acquisition of EBV and are informative for future vaccine deployment. Other infectious diseases for which MR would be similarly useful respiratory syncytial virus (RSV). A review of the putative risk factors for RSV and acute lower respiratory infections from 2015 described the huge variation between studies in how risk factors are measured, and which confounders are adjusted for. ^32^ The effect estimates in these studies were thought to be impacted substantially by confounding and the biased measurement of putative risk factors; MR has the potential to solve this issue by pinpointing which risk factors to measure and adjust for.

In an age when genetic data are widely available for an ever-growing number of risk factors and outcomes, we show MR to be a low-cost and effective way of untangling the literature surrounding the risk of acquisition of infectious conditions. Our findings demonstrate the value of MR for determining successful vaccine deployment strategies, as well as designing epidemiological studies that are appropriately adjusted for confounding.

## Data Availability

All data produced in the present study are available upon reasonable request to the authors

## Ethics approval

This analysis of secondary data was sponsored by the Academic and Clinical Central Office for Research and Development (ACCORD) of the University of Edinburgh and National Health Service Lothian, UK (AC19175). The research conducted within this study was assessed using an Usher Institute, University of Edinburgh Level 1 Ethics Self-Audit, which demonstrated that it posed no reasonably foreseeable ethical risks and so was exempt from formal ethic review by the Usher Research Ethics Group. The UK Biobank genotypic and phenotypic data used in this study were approved under application 19655.

## Author contributions

MDM, HRS, NP, and GT conceived of the work. MDM, HRS and NP designed the work. MDM and NP analysed the data for the work. All authors interpreted the data for the work. MDM drafted the work. All authors revised it critically for important content. All authors give final approval of the version to be published. All authors agree to be accountable for all aspects of the work in ensuring that questions related to the accuracy or integrity of any part of the work are appropriately investigated and resolved.

## Data availability

The source data are openly available upon application to UK Biobank using the UKBiobank data access process [http://www.ukbiobank.ac.uk/register-apply/].

## Supplementary data

Supplementary data are available at *IJE* online

## Acknowledgements

The authors wish to acknowledge Dr. Athina Spiliopoulou for useful methodological discussions and guidance.

