## Supplementary Materials for "Unpicking the Gordian knot: Mendelian randomization to elucidate the risk factors for infectious diseases, using EBV as a model pathogen"

**Supplementary Material - Unpicking the Gordian knot: Mendelian randomization to elucidate the risk factors for infe**

**Supplementary Figures:**

**Supplementary Fig 2a) Leave one out analysis for educational attainment** Educational attainment leave one out sensitivity analysis removed each instrumental variable from the Mendelian randomization, no single instrument was driving the effect estimate.	8

**Supplementary Tables:**

**Supplementary Table 1 Putative non-genetic risk factors for Epstein Barr virus infection to be explored in the Mendelian randomisation.** Six factors were identified to have a sufficient balance of evidence from Winter et al.^1^ to be considered as putative risk factors in our Mendelian randomisation analysis: total number in household, total number of sexual partners, BMI, tonsillectomy, educational attainment, and smoking status. Abbreviations: BMI – body mass index, CI – confidence intervals, EBV – Epstein Barr Virus, OR – odds ratio, Ref – references, UK- United Kingdom, USA – United States of America

| Risk Factor | Author | Year | Country | Summary | Ref |
| --- | --- | --- | --- | --- | --- |
| Total number of siblings | Levine *et al.*  Jansen *et al.* | 2012  2016 | Israel  The Netherlands | Increase in seroprevalence with increased number of siblings  EBV seropositive children had 2 or more siblings (OR= 1.35; 95% CI 1.05-1.74) | ^2,3^ |
| Total number of sexual partners | Crawford *et al.* | 2002 | UK | Prevalence of seropositivity is increased amongst those who are sexually active | ^4^ |
| BMI | Thjodleifsson *et al.*  Bertrand *et al.*  Dowd *et al.*  Spielman *et al.* | 2008  2010  2013  2014 | Iceland, Sweden, Estonia  USA  USA  USA | Lower seroprevalence with increased BMI | ^5–8^ |
| Tonsillectomy | Durovic *et al.* | 2013 |  | Lower seroprevalence amongst those with history of tonsillectomy | ^9^ |
| Educational attainment | Chen *et al.* | 2015 | Taiwan | Higher educational level associated with higher seropositivity rate | ^10^ |
| Smoking status | Levine *et al.*  Xu *et al.* | 2012  2012 | Israel  China | Seropositivity associated with smokers compared to non-smokers (OR, 3.05; 95% CI, 1.63-5.60)  Smoking associated with EBV seropositivity | ^2,6,11^ |

**Supplementary Table 2 Table of GWAS studies used in Mendelian randomization analysis.** Studies chosen for Mendelian randomization (MR) analyses. BMI, tonsillectomy, number of sexual partners, educational attainment summary statistics and instruments were extracted from the available outcomes within the TwoSampleMR package. Cigarettes per day and age at smoking initiation summary statistics were downloaded as per the publication’s instructions. Number of siblings instruments were extracted from GWAS results performed in house. BMI – body mass index, GWAS- genome wide association study.

^a^ summary statistics not from published article

| Trait | Author | Year | Sample Size |
| --- | --- | --- | --- |
| BMI | Yengo *et al.*^12^ | 2018 | 681,275 |
| Tonsillectomy | Elsworth *et al.^a^* | 2018 | 462,933 |
| Number of Sexual Partners | Elsworth *et al.^a^* | 2018 | 378,882 |
| Educational Attainment | Lee *et al.*^13^ | 2018 | 766,345 |
| Age at smoking initiation | Liu *et. al*^14^ | 2019 | 1,232,091 |
| Smoking initiation | Liu *et al.*^14^ | 2019 | 337,334 |
| Number of Siblings | In house GWAS | - | 487,409 |

| Variable | UKBiobank Cohort | | EBV serology results present and genomically deemed to be of white British ancestry (subcohort) | |
| --- | --- | --- | --- | --- |
|  | N | Column % | N | Column % |
| TOTAL | 502,616 | 100.0 | 8,244 | 100.0 |
| Age (years) | Median (IQR): 58.0 (51.0-64.0) |  | Median (IQR): 57.3 (51.0-64.0) |  |
|  | 502,614 | 99.9 | 8,244 | 100 |
| Missing data | 2 | 0.0004 | 0 | 0 |
| Sex |  |  |  |  |
| Male | 229,163 | 45.6 | 3,618 | 43.9 |
| Female | 273,453 | 54.4 | 4,626 | 56.1 |
| Total Number of Siblings | Median (IQR): 2 (1-3)  493,228 | 98.1 | Median (IQR): 2 (1-3)  8,235 | 99.9 |
| Missing data | 9,388 | 1.9 | 9 | 0.1 |
| Total Number of Sexual Partners | Median (IQR): 3 (1-6) |  | Median (IQR): 3 (1-6) |  |
| <2 | 116,685 | 23.2 | 2,019 | 24.5 |
| 2-5 | 130,739 | 26.0 | 2,186 | 26.5 |
| ≥5 | 157,745 | 31.4 | 2,616 | 31.7 |
| Missing data | 97,447 | 19.4 | 1,423 | 17.3 |
| BMI | Median (IQR): 26.7 (24.1-29.9)  499,511 | 99.4 | Median (IQR): 27.28 (24.0-29.7)  8,219 | 99.7 |
| Missing data | 3,105 | 0.6 | 25 | 0.3 |
| Tonsillectomy |  |  |  |  |
| Yes | 1,000 | 0.2 | 13 | 0.2 |
| No | 501,616 | 99.8 | 8,231 | 99.8 |
| Educational Attainment (years) | Median (IQR): 15(13-20)  492,473 | 98.0 | Median (IQR): 15 (13-20)  8,173 | 99.1 |
| Missing data | 10,143 | 2.0 | 71 | 0.9 |
| Smoking Status |  |  |  |  |
| Current | 52,885 | 10.5^a^ | 804 | 9.8 |
| Previous | 173,091 | 34.4 | 2,894 | 35.1 |
| Never | 273,588 | 54.4 | 4,511 | 54.7 |
| Missing data | 3,052 | 0.6 | 35 | 0.4 |

**Supplementary Table 3 Demographic and clinical baseline data for the UK Biobank cohort versus the sub-cohort for analysis.** Abbreviations, BMI – body mass index, EBV – Epstein-Barr Virus, IQR – interquartile range

^a^Does not add to 100% due to rounding

**Supplementary Fig 1 Manhattan plot of EBV serostatus loci**. Manhattan plot showing the strength of the associated -log10 (p value) against the chromosome location of the SNPs. Genome wide significance is represented by the red line (5x10-^8^)
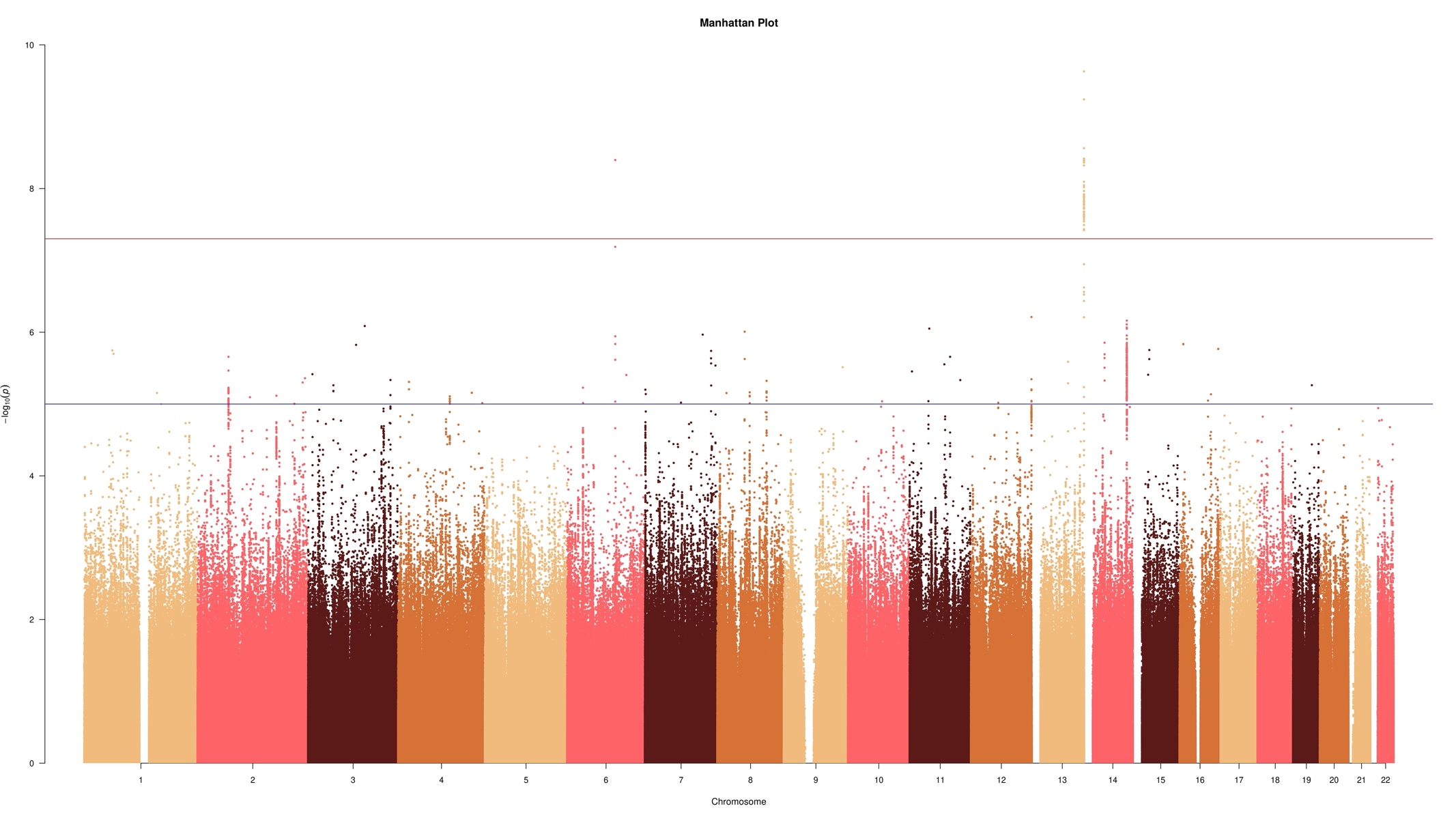

**Supplementary Table 4 – Significant GWAS hits of EBV serostatus.** Table showing the two loci that were significant after our genome wide association study (GWAS) of EBV serostatus and their nearest mapped gene. Freq- frequency.

| SNP | Chr | Start | End | Pos | A1 | A0 | Beta | SE | P Value | Freq1 | R^2^ | Nearest Gene |
| --- | --- | --- | --- | --- | --- | --- | --- | --- | --- | --- | --- | --- |
| rs1210063 | 6 | 105712424 | 105784121 | 105770008 | G | A | 0.04 | 0.007 | 4.01x10^-09^ | 0.941 | 0.004 | PREP |
| rs71449058 | 13 | 114639490 | 114882724 | 114838361 | C | T | -0.10 | 0.016 | 2.34x10^-10^ | 0.0148 | 0.005 | RASA3 |

**Supplementary Table 5 Table of heterogeneity statistics from TwoSampleMR package and number of outliers detected**. TwoSampleMR was used to test for heterogeneity for each risk factor, Outliers from each analysis were also detected and removed.

| Trait | Cochran’s Q Statistic | P Value | Outliers removed |
| --- | --- | --- | --- |
| Body mass index | 365.0263 | 0.9996492 | 28 |
| Tonsillectomy | 3.932883 | 0.7874748 | 1 |
| Total number of sexual partners | 44.02451 | 0.9124796 | 1 |
| Educational attainment | 238.3304 | 0.9866870 | 16 |
| Age at smoking initiation | 12.04674 | 0.1491306 | 0 |
| Cigarettes per day | 6.039483 | 0.5351467 | 0 |
| Number of siblings | 3.264745 | 0.3525798 | 0 |

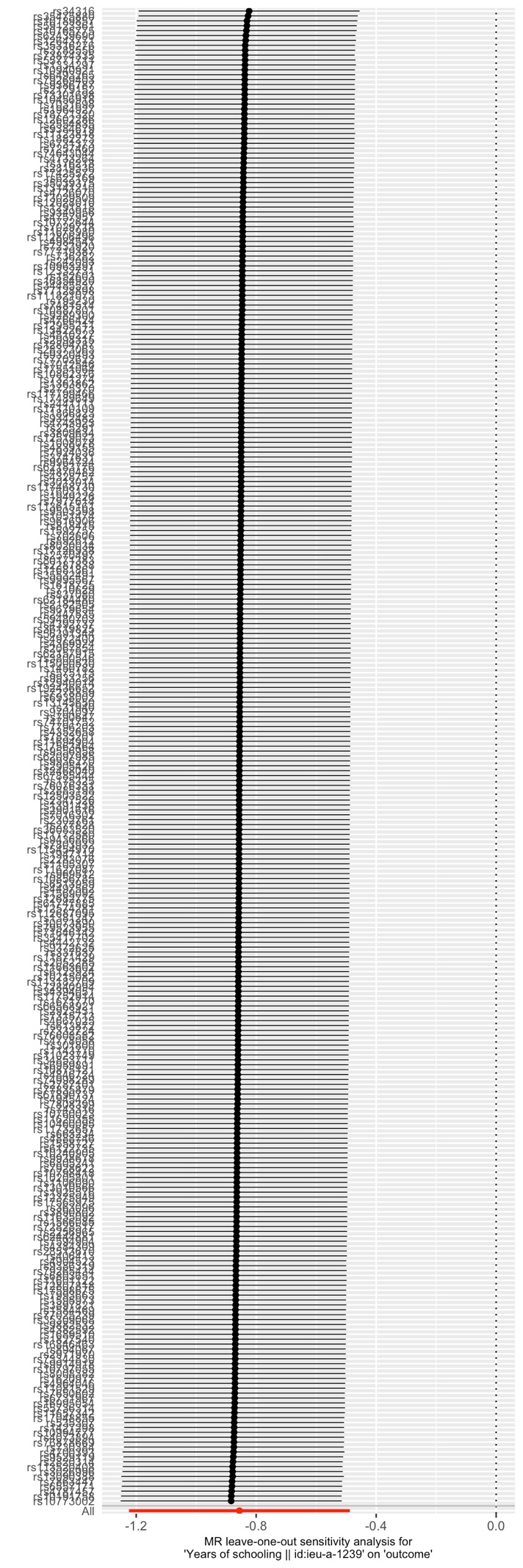

**Supplementary Fig 2a) Leave one out analysis for educational attainment.** Educational attainment leave one out sensitivity analysis removed each instrumental variable from the Mendelian randomization, no single instrument was driving the effect estimate.

**Supplementary Fig 2b) Leave one out analysis for lifetime number of sexual partners.** Sexual partners leave one out sensitivity analysis removed each instrumental variable from the Mendelian randomization, no single instrument was driving the effect estimate.

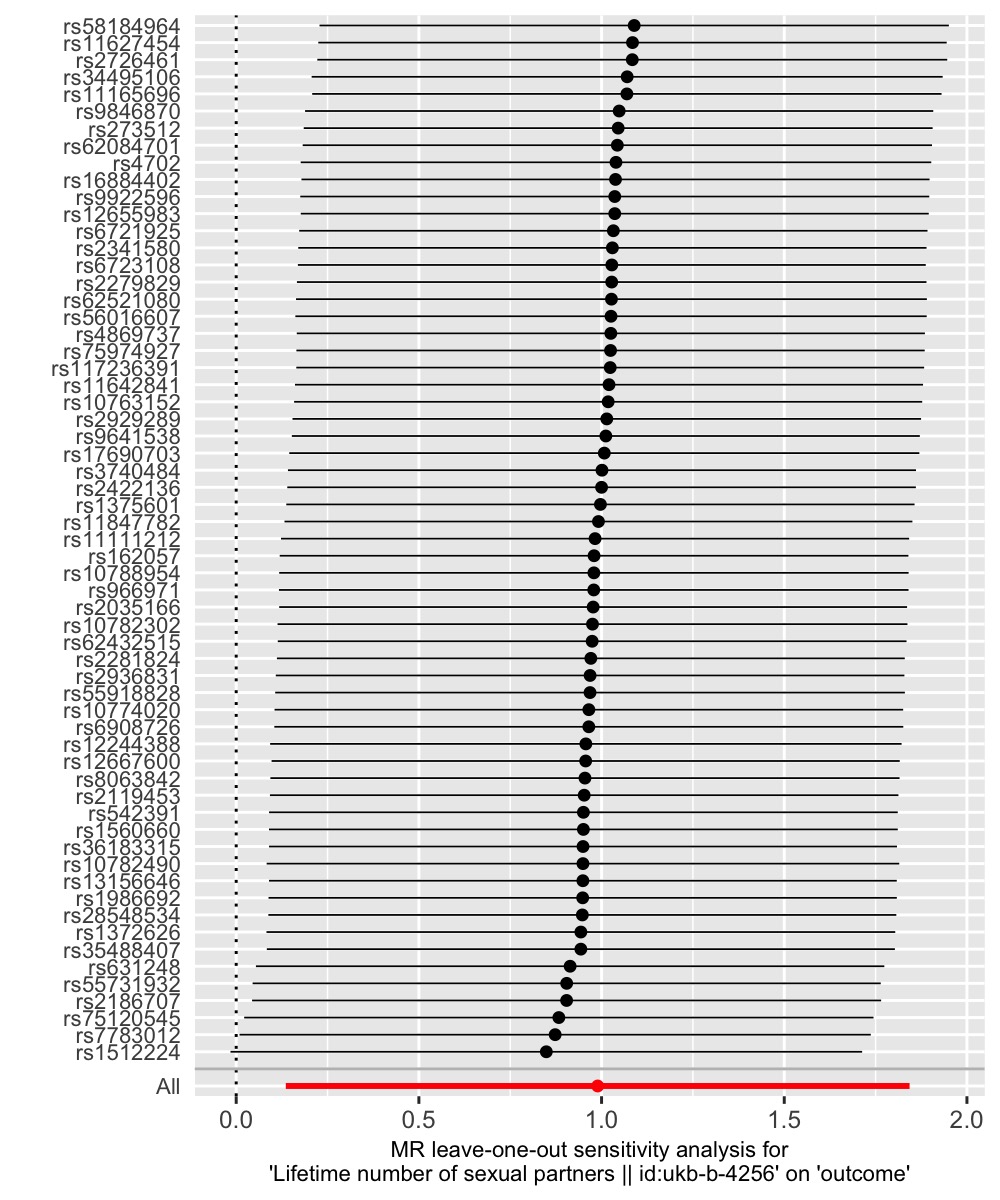

**Supplementary Fig 2c) Leave one out analysis for age at smoking initiation.** Smoking initiation leave one out sensitivity analysis removed each instrumental variable from the Mendelian randomization, no single instrument was driving the effect estimate.

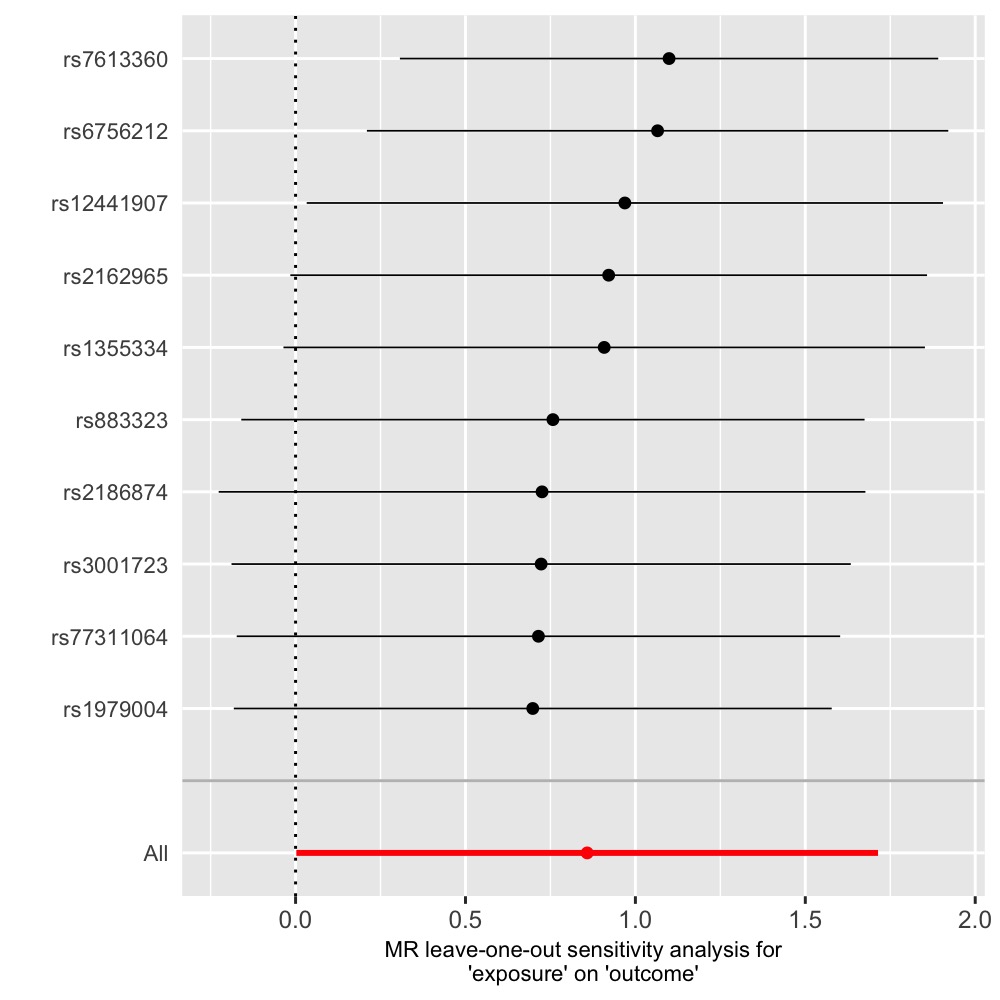
